# Development and clinical validation of blood-based multibiomarker models for the evaluation of brain amyloid pathology

**DOI:** 10.1101/2025.02.27.25322892

**Authors:** Darren M. Weber, Matthew A. Stroh, Steven W. Taylor, Robert J. Lagier, Judy Z. Louie, Nigel J. Clarke, David E. Vaillancourt, Sruti Rayaprolu, Ranjan Duara, Michael K. Racke

## Abstract

**Background and Objectives:** Plasma biomarkers provide new tools to evaluate patients with mild cognitive impairment (MCI) for Alzheimer’s disease (AD) pathology. Such tools are needed for anti-amyloid therapies that require efficient and accurate diagnostic evaluation to identify potential treatment candidates. This study sought to develop and evaluate the clinical performance of a multi-marker combination of plasma beta-amyloid 42/40 (Aβ42/40), ptau-217, and *APOE* genotype to predict amyloid PET positivity in a diverse cohort of patients at a memory clinic and evaluate >4,000 results from “real-world” specimens submitted for high-throughput clinical testing.

**Methods:** Study participants were from the 1Florida AD Research Center (ADRC). Demographics, clinical evaluations, and amyloid PET scan data were provided with plasma specimens for model development for the intended-use cohort (MCI/AD: n=215). Aβ42/40 and ApoE4 proteotype (reflecting high-risk *APOE* L4 alleles) were measured by mass spectrometry and ptau-217 by immunoassay. A likelihood score model was determined for each biomarker separately and in combination. Model performance was optimized using 2 cutpoints, 1 for high and 1 for low likelihood of PET positivity, to attain ≥90% specificity and sensitivity. These cutpoints were applied to categorize 4,326 real-world specimens and an expanded cohort stratified by cognitive status (normal cognition [NC], MCI, AD).

**Results:** For the intended-use cohort (46.0% prevalence of PET-positivity), a combination of Aβ42/40, ptau-217, and *APOE4* allele count provided the best model with a receiver operating characteristic area under the curve (ROC-AUC) of 0.942 and with 2 cutpoints fixed at 91% sensitivity and 91% specificity yielding a high cutpoint with 88% positive predictive value (PPV) and 87% accuracy and a low cutpoint with 91% negative predictive value (NPV) and 85% accuracy. Incorporating *APOE4* allele count also reduced the percentage of patients with indeterminate risk from 15% to 10%. The cutpoints categorized the real-world clinical specimens as having 42% high, 51% low, and 7% indeterminate likelihood for PET positivity and differentiated between NC, MCI, and AD dementia cognitive status in the expanded cohort.

**Discussion:** Combining plasma biomarkers Aβ42/40, ptau-217, and *APOE4* allele count is a scalable approach for evaluating patients with MCI for suspected AD pathology.

**Key Takeaways:**

1. The approval of disease-modifying therapies for Alzheimer’s disease ushers in the need for accessible, affordable, and accurate blood-based testing for Alzheimer’s pathology.
2. Models implementing multiple analytes have demonstrated high performance in identifying patients with brain amyloid pathology.
3. We developed high-throughput, robust, multiple-analyte assays and models aimed at predicting the likelihood of amyloid PET positivity.
4. We report two models with excellent performance in alignment with current recommendations for blood-based testing.
5. Aβ42/40 + ptau-217 + *APOE4* allele count provided the best prediction for amyloid PET positivity when sensitivity and specificity were both fixed at 91%.

## 1. INTRODUCTION

Alzheimer’s disease (AD) is a progressive neurodegenerative disorder characterized by beta-amyloid (Aβ) plaques and neurofibrillary tangles in the brain that are associated with cognitive impairment.^1^ Previously, PET imaging ^2^ or CSF evaluation were the only available approaches to determine levels of Aβ and phosphorylated tau protein^3^ (ptau, a marker of neurofibrillary tangles) for diagnosis of AD pathology. However, these methods are perceived as invasive and/or costly. Recently approved disease-modifying monoclonal antibody treatments for AD target specific soluble aggregates of Aβ^4,5^ and are indicated for individuals with mild cognitive impairment (MCI) or mild AD. Eligibility for these treatments requires confirmation of amyloid pathology and *APOE* L4 allele (*APOE4*) carrier status prior to administration. Scalable diagnostic tests that are accurate, less invasive, and affordable can fulfill this need.^6^

Recent advances in blood-based biomarkers (BBM) provide clinicians with a more cost-effective and accessible means to evaluate biomarkers such as Aβ and ptau.^7^ High-throughput mass spectrometry (MS) and immunoassay platforms combined with automated sample preparation provide a scalable approach to measure large numbers of blood specimens with fast turn-around times. Recently, we developed a liquid chromatography-tandem MS (LC-MS/MS) plasma test to evaluate cerebral Aβ pathology using a ratio of peptides beta-amyloid 42 (Aβ42) to beta-amyloid 40 (Aβ40) (ie, Aβ42/40) to predict amyloid PET status.^8,9^ In addition, plasma tau protein phosphorylated at threonine 217 (ptau-217) is another excellent marker of AD-related neuropathology.^10^

The Alzheimer’s Association recently included Aβ42 and ptau-217 in their guidelines as BBMs for core AD pathologies and processes.^11^ Furthermore, combinations of plasma biomarkers can provide greater accuracy in identifying and differentiating AD dementia from other neurodegenerative disorders.^12–16^ Additionally, some studies demonstrated that inclusion of *APOE4* allele carrier status or allele count, which confer elevated risk for AD, can improve model performance when combined with the plasma Aβ42/40 ratio and/or ptau-217,^15,17^ while others showed no statistically significant improvement.^8,14^

For combinations of plasma biomarkers to have substantial clinical value, they should have acceptable diagnostic performance on par with currently accepted CSF tests.^18^ In addition, BBMs should have high test-retest reliability and scalability for high throughput to be considered useful for clinical implementation^19,20^ as cost-effective diagnostic tests to identify potential candidates for anti-amyloid therapies.^16,21^

In the current study, our objective was to develop a multibiomarker combination that (1) achieved acceptable performance metrics while maintaining sensitivities and specificities ≥90% using specimens from a well-characterized cohort of patients with MCI and AD; and (2) uses robust high-throughput assays based on accessible cost-effective, low-resolution LC-MS/MS and immunoassay methods.^9,16^

We note that these recommendations have been met previously using ptau-217 alone, but in a racially (97% white) and ethnically homogeneous cohort with a high prevalence (64%) of amyloid positivity^16^ and with a model developed using a combination of ptau-217/np-tau217 and Aβ42/40 (94% white and 53% prevalence of amyloid positivity) on a high-resolution MS-based platform.^14^

In our cohort, patients were age-, sex-, ethnicity-, and years-of-education matched as PET-negative (n=116) and PET-positive (n=99), representing a lower prevalence (46%) of PET positivity and greater ethnic diversity (54% Hispanic) than prior studies. In addition to using plasma-based Aβ42/40 and ptau-217, we examined the effect of adding *APOE*4 genotype information (ie, carrier or noncarrier vs allele count, ie, 0, 1, or 2 alleles) to the combination of markers. Ultimately, we aim to assess the usefulness of these tests to effectively identify candidates for anti-amyloid therapy.

## 2. METHODS

### 2.1 Study Design and clinical evaluation

This cross-sectional study included plasma specimens from 215 participants in the intended-use cohort enrolled with 1Florida Alzheimer’s Disease Research Center (ADRC). All participants underwent amyloid PET imaging as well as cognitive testing and included 170 participants with MCI and 45 participants with AD (**Table 1**). Participants had a minimum of a 6th grade reading level and spoke English and/or Spanish as their primary language. Participant ethnicity was self-reported.

**Table 1:**
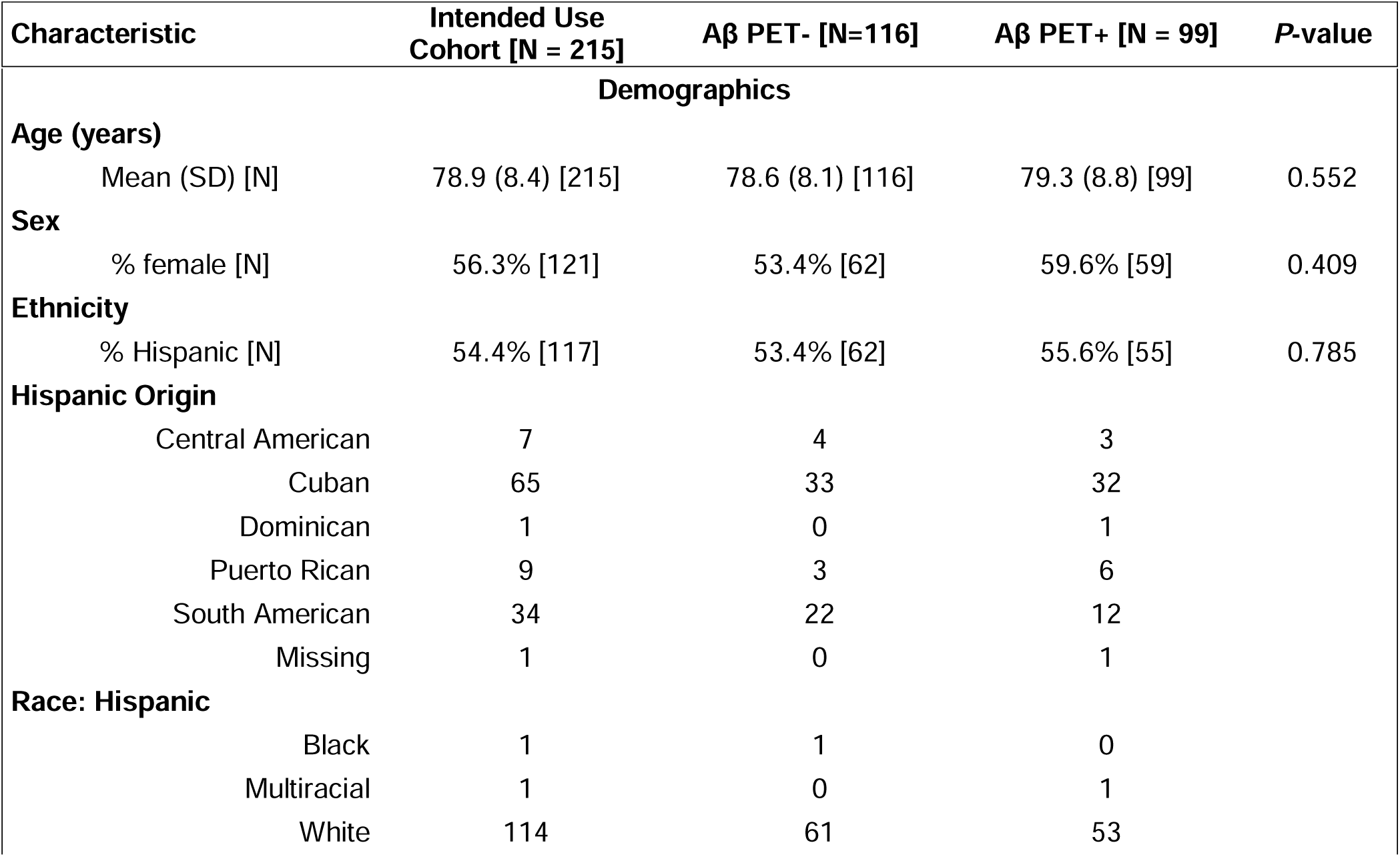
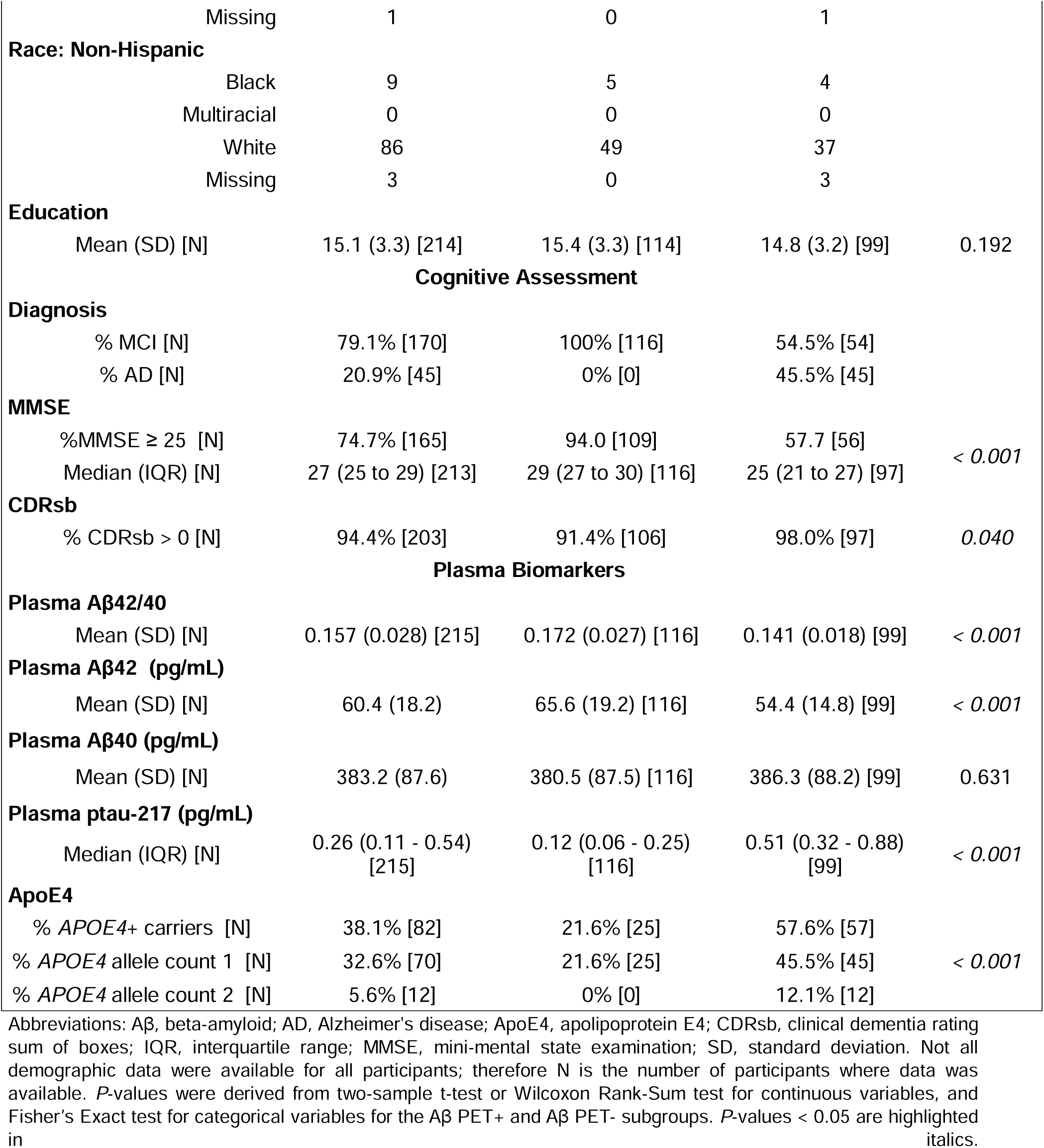
Baseline characteristics by amyloid PET status.

| Characteristic | Intended Use Cohort [N = 215] | Aβ PET- [N=116] | Aβ PET+ [N = 99] | P-value |
| --- | --- | --- | --- | --- |
| **Demographics** |  |  |  |  |
| **Age (years)** |  |  |  |  |
| Mean (SD) [N] | 78.9 (8.4) [215] | 78.6 (8.1) [116] | 79.3 (8.8) [99] | 0.552 |
| **Sex** |  |  |  |  |
| % female [N] | 56.3% [121] | 53.4% [62] | 59.6% [59] | 0.409 |
| **Ethnicity** |  |  |  |  |
| % Hispanic [N] | 54.4% [117] | 53.4% [62] | 55.6% [55] | 0.785 |
| **Hispanic Origin** |  |  |  |  |
| Central American | 7 | 4 | 3 |  |
| Cuban | 65 | 33 | 32 |  |
| Dominican | 1 | 0 | 1 |  |
| Puerto Rican | 9 | 3 | 6 |  |
| South American | 34 | 22 | 12 |  |
| Missing | 1 | 0 | 1 |  |
| **Race: Hispanic** |  |  |  |  |
| Black | 1 | 1 | 0 |  |
| Multiracial | 1 | 0 | 1 |  |
| White | 114 | 61 | 53 |  |

|  |  |  |  |  |  |
| --- | --- | --- | --- | --- | --- |
|  | Missing | 1 | 0 | 1 |  |
| Race: Non-Hispanic |  |  |  |  |  |
|  | Black | 9 | 5 | 4 |  |
|  | Multiracial | 0 | 0 | 0 |  |
|  | White | 86 | 49 | 37 |  |
|  | Missing | 3 | 0 | 3 |  |
| Education |  |  |  |  |  |
|  | Mean (SD) [N] | 15.1 (3.3) [214] | 15.4 (3.3) [114] | 14.8 (3.2) [99] | 0.192 |
| Cognitive Assessment |  |  |  |  |  |
| Diagnosis |  |  |  |  |  |
|  | % MCI [N] | 79.1% [170] | 100% [116] | 54.5% [54] |  |
|  | % AD [N] | 20.9% [45] | 0% [0] | 45.5% [45] |  |
| MMSE |  |  |  |  |  |
|  | %MMSE ≥ 25 [N] | 74.7% [165] | 94.0 [109] | 57.7 [56] | < 0.001 |
|  | Median (IQR) [N] | 27 (25 to 29) [213] | 29 (27 to 30) [116] | 25 (21 to 27) [97] |  |
| CDRsb |  |  |  |  |  |
|  | % CDRsb > 0 [N] | 94.4% [203] | 91.4% [106] | 98.0% [97] | 0.040 |
| Plasma Biomarkers |  |  |  |  |  |
| Plasma Aβ42/40 |  |  |  |  |  |
|  | Mean (SD) [N] | 0.157 (0.028) [215] | 0.172 (0.027) [116] | 0.141 (0.018) [99] | < 0.001 |
| Plasma Aβ42 (pg/mL) |  |  |  |  |  |
|  | Mean (SD) [N] | 60.4 (18.2) | 65.6 (19.2) [116] | 54.4 (14.8) [99] | < 0.001 |
| Plasma Aβ40 (pg/mL) |  |  |  |  |  |
|  | Mean (SD) [N] | 383.2 (87.6) | 380.5 (87.5) [116] | 386.3 (88.2) [99] | 0.631 |
| Plasma ptau-217 (pg/mL) |  |  |  |  |  |
|  | Median (IQR) [N] | 0.26 (0.11 - 0.54) [215] | 0.12 (0.06 - 0.25) [116] | 0.51 (0.32 - 0.88) [99] | < 0.001 |
| ApoE4 |  |  |  |  |  |
|  | % APOE4+ carriers [N] | 38.1% [82] | 21.6% [25] | 57.6% [57] |  |
|  | % APOE4 allele count 1 [N] | 32.6% [70] | 21.6% [25] | 45.5% [45] | < 0.001 |
|  | % APOE4 allele count 2 [N] | 5.6% [12] | 0% [0] | 12.1% [12] |  |

Abbreviations: Aβ , beta-amyloid; AD, Alzheimer's disease; ApoE4, apolipoprotein E4; CDRsb, clinical dementia rating sum of boxes; IQR, interquartile range; MMSE, mini-mental state examination; SD, standard deviation. Not all demographic data were available for all participants; therefore N is the number of participants where data was available. *P*-values were derived from two-sample t-test or Wilcoxon Rank-Sum test for continuous variables, and Fisher's Exact test for categorical variables for the Aβ PET+ and Aβ PET- subgroups. *P*-values < 0.05 are highlighted in italics.

Participants underwent an extensive neurological and neuropsychological evaluation, including the clinical dementia rating sum of boxes scale (CDRsb)^22^ and the mini-mental state examination (MMSE).^23^ Diagnoses of MCI and AD dementia were determined using a previously described algorithmic procedure,^24^ which combines the neuropsychological diagnosis with the CDRsb scores.

To evaluate the diagnostic performance of the biomarkers for identifying cognitive status, we evaluated and compared additional specimens from 56 participants in the same 1Florida ADRC cohort who were assessed to have normal cognition (NC; 49 PET-negative/7 PET-positive) (**eTable 1**).

In addition, we performed a retrospective analysis of 4,326 consecutive plasma specimens submitted to Quest Diagnostics for plasma Aβ42/40, ptau-217, and ApoE proteoform analysis testing. This was a limited dataset (as defined by https://www.hhs.gov/hipaa/for-professionals/special-topics/research/index.html) with only patient age and sex information available (**eTable 2)**.

### 2.2 Amyloid PET imaging

Amyloid PET imaging was carried out using [^18^F]florbetaben or [^18^F]florbetapir tracers on a Siemens 16 Biograph PET/CT scanner, as described previously.^25^ Global amyloid status (+/-) was determined based on visual read by an experienced rater who was blinded to diagnosis and cognitive status.^25^ All participants underwent an amyloid PET scan within 12 months of plasma collection.

### 2.3 Plasma biomarker analysis

Blood specimens were collected by venipuncture into 10-mL tubes containing EDTA as anticoagulant and kept on ice (<1 h) until centrifugation at 1,200 relative centrifuge force for 12 min at room temperature. Plasma aliquots (0.5 mL) were transferred into polypropylene tubes and stored at −80 °C until analysis. All plasma specimens were deidentified, and results were blinded during the analysis.

#### 2.3.1 Plasma A**β**42/40 ratio and ApoE proteoform assays

The LC-MS/MS-based assays for the plasma Aβ42/40 ratio and ApoE proteoforms have been described in detail elsewhere.^9^ All sample preparation steps were fully automated using a Hamilton Star liquid handler (Hamilton, Reno, NV). LC-MS/MS was performed using a Transcend Vanquish TLX-4 TurboFlow UPLC, with a high-throughput 4-LC column configuration, coupled to a TSQ Altis Plus Triple Quadrupole MS (both Thermo Fisher Scientific, Waltham, MA); data were analyzed using TraceFinder Clinical Research v5.1 software (Thermo Fisher Scientific).

For the Aβ42/40 ratio, Aβ40 and Aβ42 were enriched by immunoprecipitation from plasma (0.5 mL), digested with Lys-C, and desalted and concentrated by solid phase extraction. Characteristic peptides for Aβ40 (Aβ29-40) and Aβ42 (Aβ29-42) were chromatographically separated, detected, and quantified by LC-MS/MS.^9^

For ApoE proteoform determination, plasma specimens (25 µL) were reduced, alkylated, and proteolytically digested with trypsin. Unique tryptic peptides to the ApoE2 and ApoE4 proteoforms, as well as shared peptides between the ApoE2 and ApoE3 proteoforms, and the ApoE3 and ApoE4 proteoforms, were chromatographically separated and detected by LC-MS/MS. ApoE proteoforms were determined by the presence or absence of the unique and shared peptides and used to establish ApoE proteotype, which has previously been shown to be 100% concordant with *APOE* genotype.^9^

Both assays were validated according to Clinical Laboratory Improvement Amendments (CLIA) guidance. For the Aβ42/40 ratio, assay imprecision (random error) and bias (technical error) was measured previously to establish the technical robustness^8^ of the test per Rabe et al.^26^ Total error was calculated as described in section 2.4 Statistical analysis.

#### 2.3.2 Plasma ptau-217 assay

On the day of the assay, frozen calibrators, quality control samples (QCs), and plasma specimens were thawed and brought to room temperature. Plasma specimens were centrifuged to remove any debris, and plasma ptau-217 levels were quantified using the Lumipulse G ptau-217 - Plasma Immunoreaction Cartridges RUO (for research use only) assay kit on a Lumipulse G System (Fujirebio Inc, Malvern, PA), which is a fully automated chemiluminescent enzyme immunoassay (CLEIA) platform.

The assay was validated according to CLIA guidance and data were used to evaluate technical robustness per Rabe et al.^26^ Inter-assay imprecision (random error) was determined by taking the average of 2 QCs analyzed in duplicate over 14 nonconsecutive days. Bias (technical error) was experimentally determined by analyzing 100 previously assayed plasma specimens using a different lot of reagents. Values for ptau-217 were compared as a percentile difference calculated between the mean values of the original results and retest results, as has been reported elsewhere.^8,26^ Total error was calculated as described in Section 2.4.3.

### 2.4 Statistical analysis

#### 2.4.1 Demographic, clinical, and biomarker comparisons

Participant demographics were summarized for the full cohort and further stratified by amyloid PET status. Continuous variables were summarized as mean and standard deviation (SD) or median and interquartile range. Categorical variables were summarized as percent and count (N). Tests for difference between PET status were assessed by 2-sample t-test or Wilcoxon Rank-Sum test for continuous variables and Fisher’s Exact test for categorical variables. Significance was determined with a threshold of *P*≤.05.

#### 2.4.2 Logistic regression

Logistic regression modeling was employed to assess the association between predictor variables and amyloid PET status to estimate model linear predictors. The ptau-217 variable as a predictor in the models was natural log transformed.^27^ Akaike Information Criterion (AIC) was used to select the final model. Amyloid PET positivity likelihood scores in probability scale were calculated by transforming model linear predictors in natural exponential function. Tests for differences in the distribution of amyloid PET positivity likelihood scores between amyloid PET status were assessed by Wilcoxon Rank-sum test. Model performances of predictors in predicting PET positivity were evaluated by ROC curve analysis. Differences between ROC curves were assessed by the method of DeLong.^28^ Comparisons of probability/likelihood scores when stratified by amyloid PET status were conducted using a two-tailed paired t-test.

In addition, model performances were also evaluated by a 2-cutpoint approach with the lower cutpoint establishing maximized sensitivity and the upper cutpoint establishing maximized specificity. PPV and NPV were calculated at each classification threshold via Bayes’ rule, with estimated sensitivity, specificity, and prevalence as inputs. Recommended performance criteria stipulate that those patients who fall in the indeterminate zone between the two cutpoints represent ≤20% of the population.^18^

#### 2.4.3 Technical robustness

To investigate the effects of total error (assay imprecision+potential bias) for the Aβ40, Aβ42, and ptau-217 markers on the predictive performance of the best model, data simulations were performed. Estimates for analytical imprecision and bias were previously established for each marker (data not included). Based on those error estimates, Monte Carlo simulations were performed to generate 10,000 replicate study data sets (N=215) with randomly assigned errors drawn from a Gaussian distribution (mean=0, %CV=6%) and fixed bias offsets added to each biomarker value. Aβ42/40 ratios were generated for each paired set of perturbed amyloid markers. For each simulated data set, observations were scored by the best model (each observation retained its original *APOE4* allele count for scoring) and assigned a new predicted PET+ class (low, indeterminate, or high likelihood) based on the established classification thresholds. Predicted PET+ classes for each simulated data set were tabulated against the original (non-perturbed) data and reclassification rates were estimated. The average (95% CI) reclassification rate due to overall assay error was estimated as the median (2.5^th^ percentile, 97.5^th^ percentile) of the 10,000 simulated reclassification rates.

All data analyses were performed using R version 4.3.1 (The R Foundation for Statistical Computing).

### 2.5 Data availability

The raw data supporting the conclusions of this article will be made available by the authors on request.

### 2.6 Ethics statement

The studies involving humans were approved by Mount Sinai Medical Center IRB. The studies were conducted in accordance with the local legislation and institutional requirements. The human samples used in this study were acquired as part of a previous study for which ethical approval was obtained. Written informed consent for participation from the participants or the participants’ legal guardians/next of kin in accordance with the national legislation and institutional requirements.

## 3. RESULTS

### 3.1 Demographic, clinical, and biomarker comparisons

The mean age of participants in the intended-use cohort was 79 years (SD, 8 years; range 52-97) with almost 80% having MCI and the remaining having AD dementia (**Table 1**). Women (56%) were slightly more represented than men and there was a slight majority (54%) of participants with Hispanic ethnicity (56% of these participants were of Cuban origin, **Table 1**). Most participants had a CDRsb score >0 (94%) and an MMSE score ≥25 (nearly 75%, **Table 1**).

Compared to PET-negative participants, PET-positive participants had a significantly higher percentage with a CDRsb score >0 (*P*=.040) and significantly lower median MMSE scores (*P*<.001, **Table 1**). No significant differences between patient age, sex, years-of-education, and ethnicity were found between those who were PET-negative and PET-positive.

Compared to PET-negative participants, PET-positive participants had significantly lower mean levels of plasma Aβ42, higher median levels of ptau-217, and a significantly lower mean Aβ42/40 ratio (*P*<.001 for all, **Table 1**). No difference in plasma Aβ40 levels between PET-positive and PET-negative groups was observed.

### 3.2 Prediction of amyloid PET status

As described below, the individual and multi-marker prediction of amyloid PET status using either single or 2-cutpoint approaches were evaluated in terms of optimizing test performance and to meet recommended 90% sensitivity/90% specificity criteria for confirmatory testing.^18^ For the 2-cutpoint approach, this included keeping the percentage of patients classified as having indeterminate likelihood of PET positivity ≤20%.^18^

#### 3.2.1 Diagnostic performance of individual and combined BBMs for predicting amyloid PET status using a single cutpoint

ROC-AUCs for PET prediction using Aβ42/40 alone (0.851, 95%CI 0.796 to 0.905) and ptau-217 alone (0.846, 95%CI 0.808 to 0.905) significantly improved (*P*<.001) when they were combined (0.929, 95%CI 0.894 to 0.963) (**eTable 3, Figure 1**). The AIC for the model that combined Aβ42/40 and ptau-217 predictors was 195, lower than Aβ42/40 (AIC = 256) and ptau-217 (AIC = 263) alone (**eTable 3)**. Further improvement on amyloid PET prediction was achieved when *APOE4* results were added to the model, with *APOE4* carrier status (positive/negative) yielding an AUC of 0.938 (95%CI 0.906 to 0.969), and *APOE4* allele count yielding an AUC of 0.942 (95%CI 0.912 to 0.972). However, only the *APOE4* allele count model AUC (AIC = 186, *P*=0.033), and not the carrier status model (AIC = 189, *P* =0.177), reached statistical significance when compared to the model without *APOE4* results added. Despite high AUCs for each combined BBM predictor, none of the models achieved ≥90% sensitivity/specificity when using a single cutpoint (**eTable 3**, **Figure 1**).

**Figure 1:**
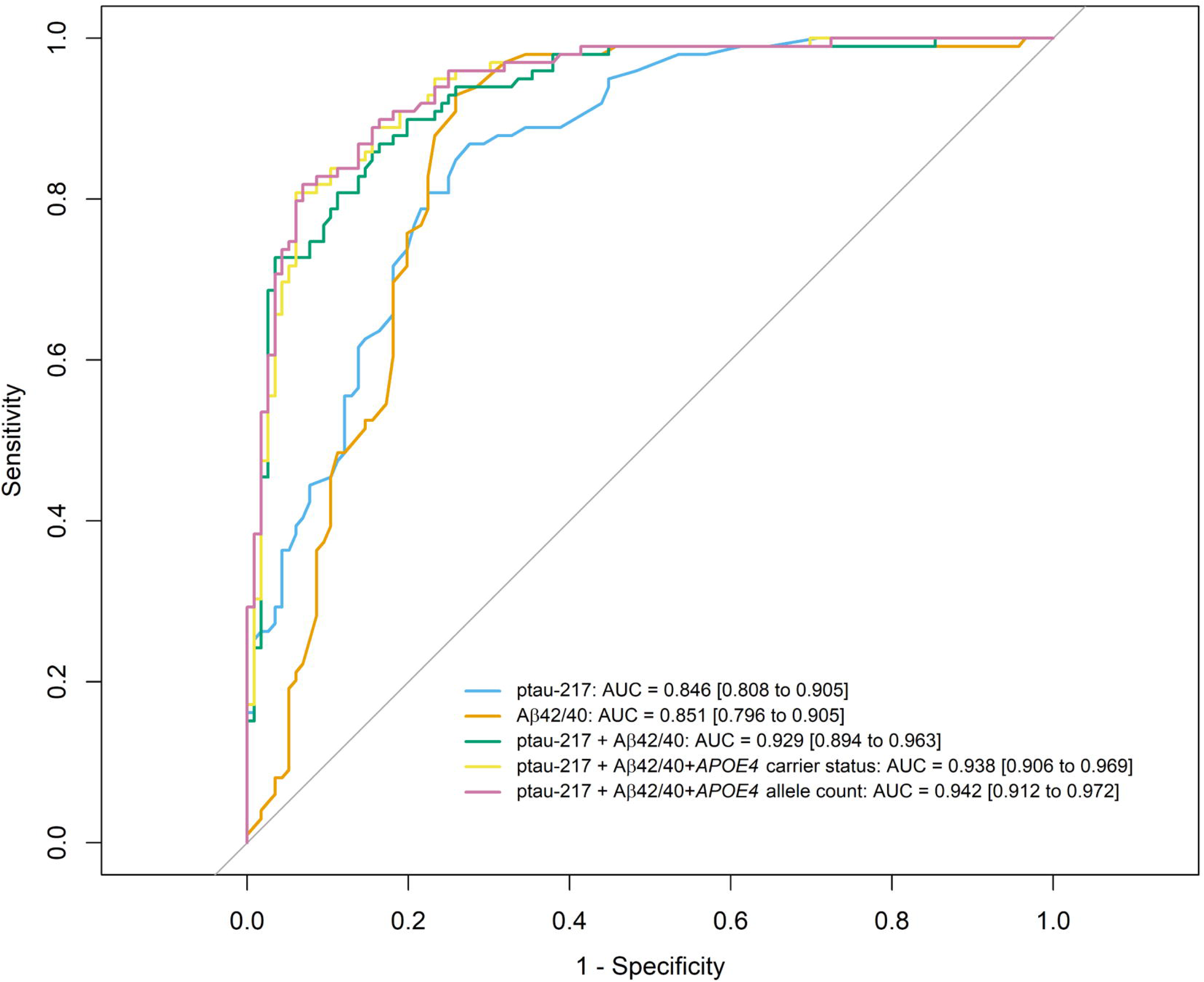
Receiver operating characteristic-area under the curve (ROC-AUC) analysis for individual and combined biomarkers for prediction of PET status. Abbreviations: Aβ, beta-amyloid; *APOE4*, APOE ε4; AUC, area under the curve.

#### 3.2.2 Diagnostic performance of individual and combined BBMs for predicting amyloid PET status using a 2-cutpoint method

At probability cutpoints establishing ≥90% sensitivity and specificity, no individual biomarker met the criterion for ≤20% indeterminate classifications.^18^ Aβ42/40 classified 34% and ptau-217 classified 42% of the intended-use population as having an indeterminate result. Combining Aβ42/40 and ptau-217 reduced the indeterminate results to 15%, and further addition of *APOE4* allele count reduced them to 10% (**Figure 2**, **Table 2**). The reduction represented 21 of 32 participants (66%) being reclassified in the correct direction relative to their PET status (wrong classification to indeterminate [n=6] or indeterminate to correct classification [n=15]). The distribution of results for the intended-use population for the Aβ42/40+ptau-217+*APOE4* allele count model is summarized in **Figure 3A**.

**Figure 2:**
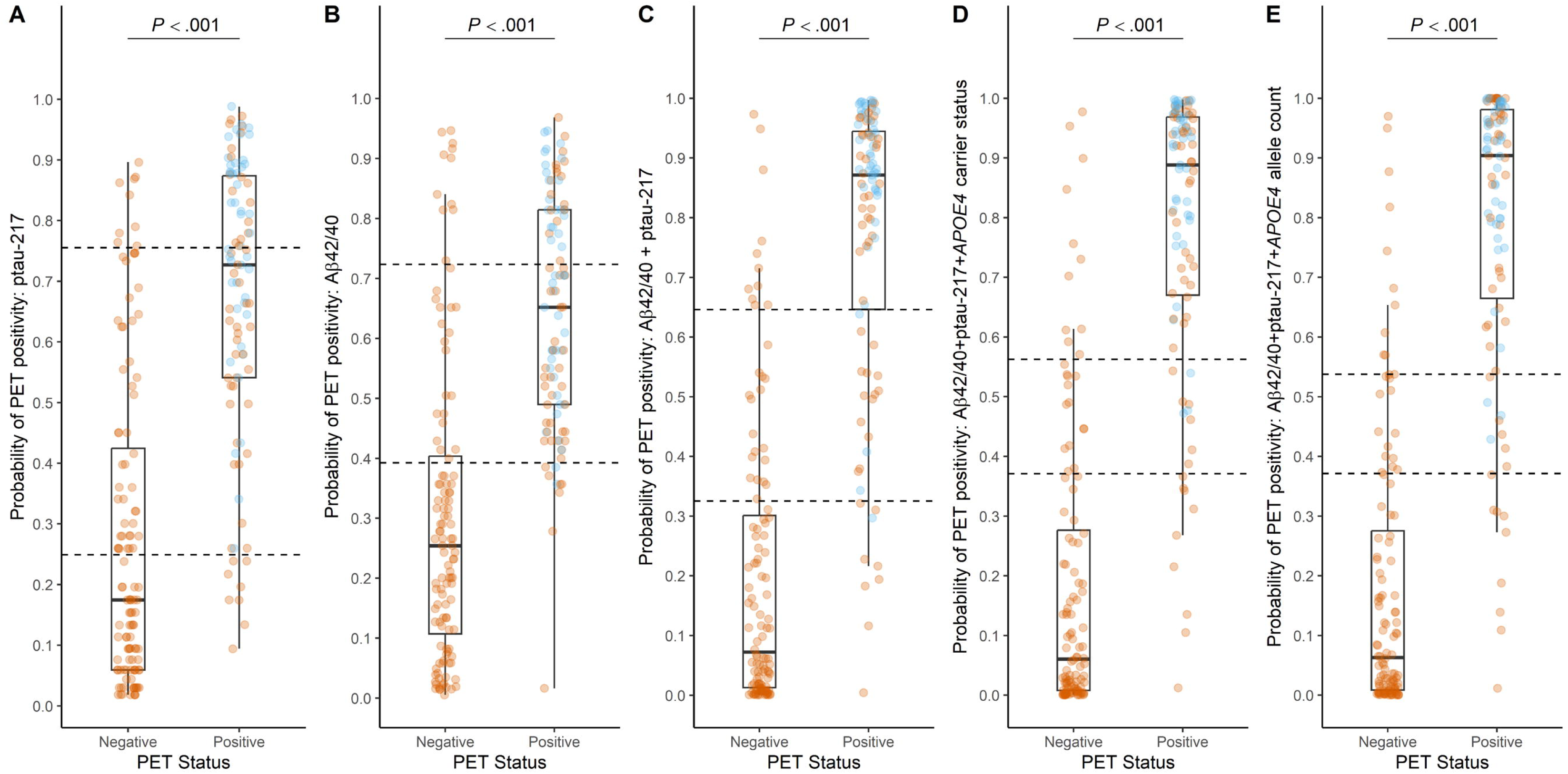
Boxplots of likelihood scores stratified by PET status for (A) ptau-217, (B) Aβ42/40, (C) Aβ42/40 + ptau-217, (D) Aβ42/40 + ptau-217 +*APOE4* carrier status, and (E) Aβ42/40 + ptau-217 + *APOE4* allele count. Orange dots represent individuals that are MCI; blue dots represent individuals that are AD. Horizontal dashed lines indicate high and low likelihood cutoffs. Results between these lines indicate neither a low nor high likelihood (ie, an indeterminate result).

**Figure 3:**
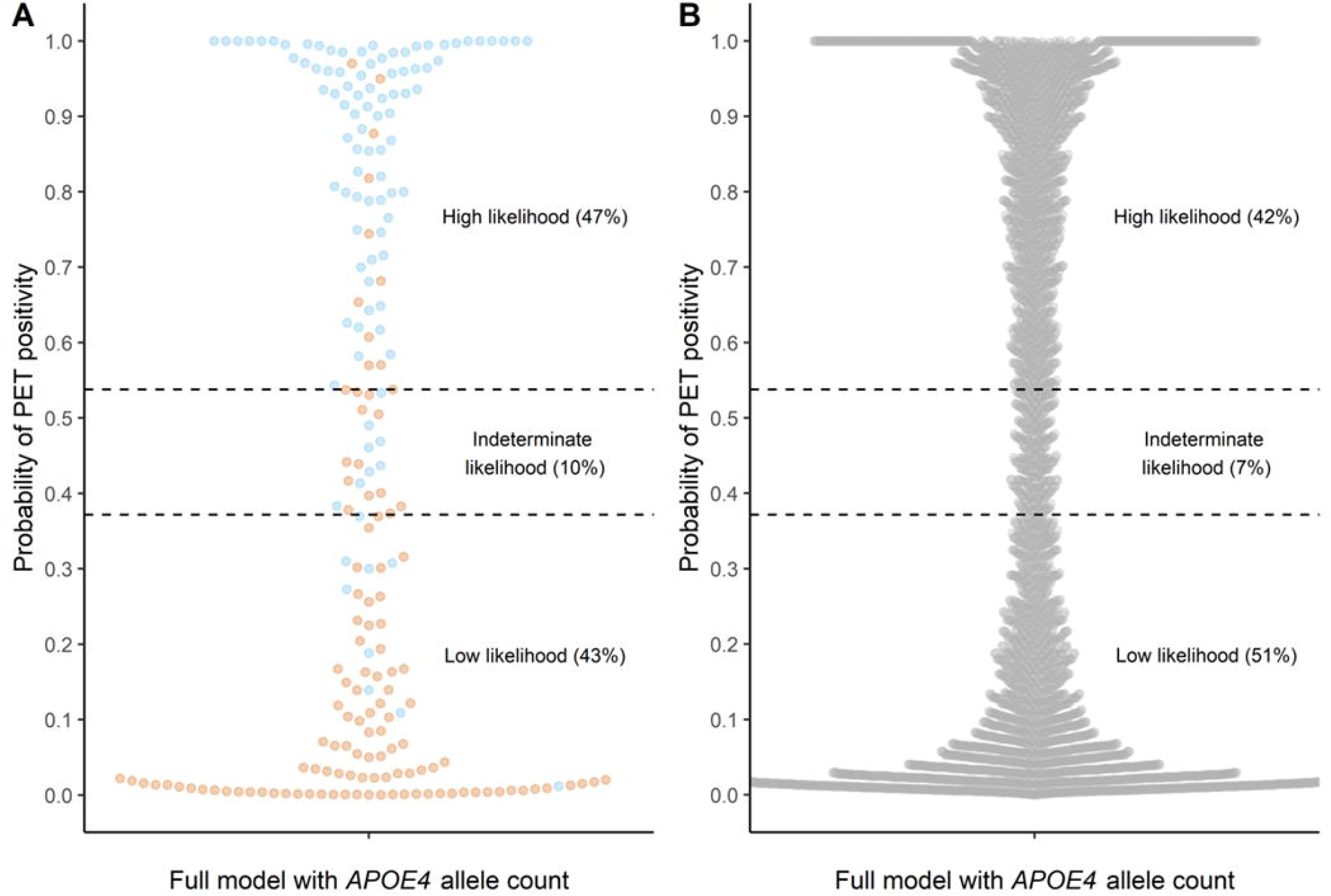
Distribution of model results for A) the intended use population, and B) real-world clinical specimens. Orange dots represent individuals that are amyloid PET negative; blue dots represent those that are amyloid PET positive in panel A. Grey dots represent individuals whose amyloid PET status is unknown. Horizontal dashed lines indicate high and low likelihood cutoffs.

**Table 2:**
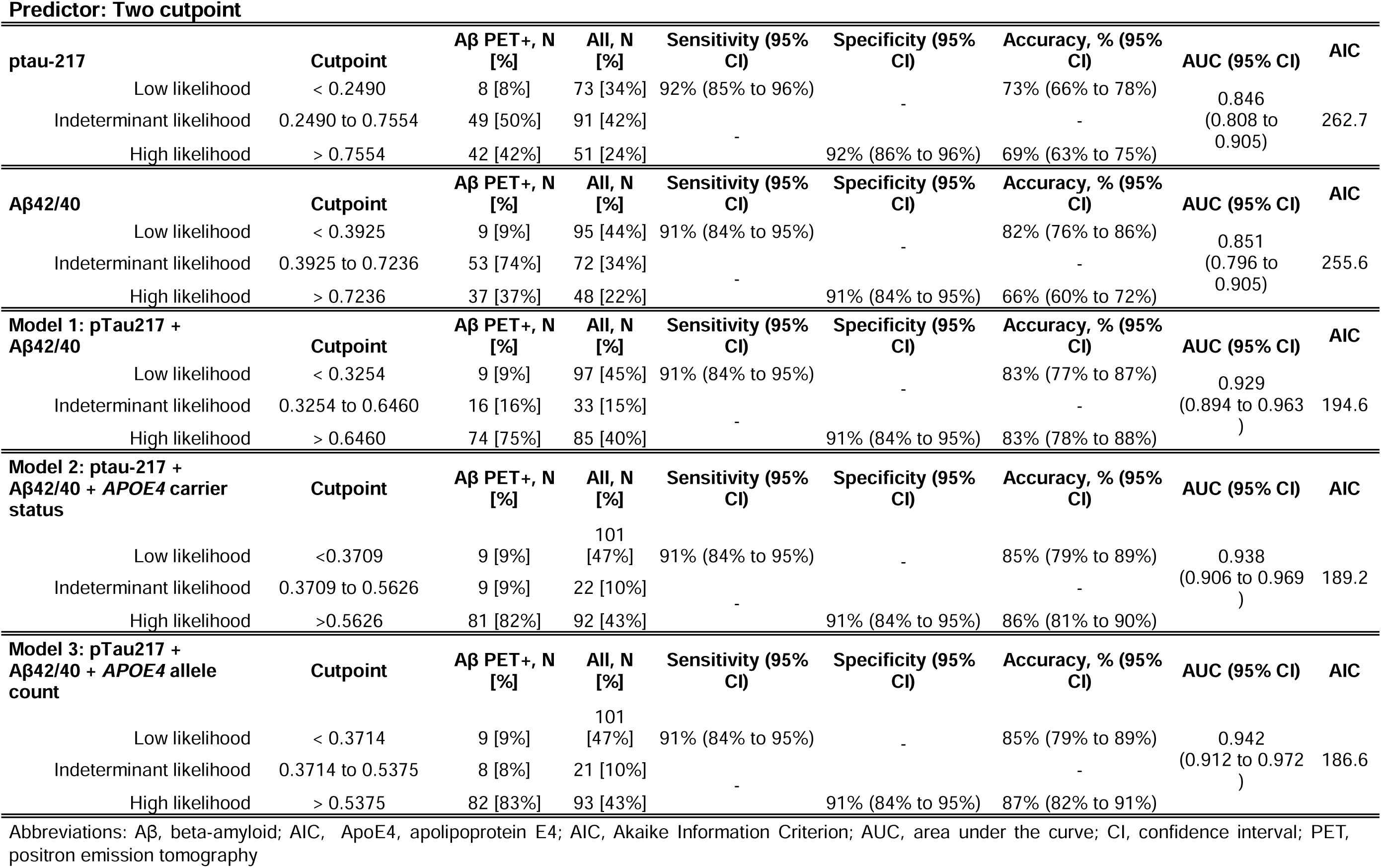
Model performance for prediction of amyloid PET status at a defined sensitivity and specificity.

The models with vs without *APOE4* allele count with the 2-cutpoints set for 91% sensitivity and 91% specificity at the study prevalence of 46% of PET-positivity were compared. PPVs were 88% with *APOE4* allele count and 87% without and NPVs were 91% for both models (**Table 3**). At 50% prevalence, which is the prevalence expected in an MCI population,^29^ the estimated PPV and NPV would both reach 90% for the model with *APOE4* allele count compared to 89% for both PPV and NPV for the model without. At lower prevalence of PET-positivity, such as in a primary care setting where the prevalence would be closer to 20%,^30^ the model predictors with and without *APOE4* allele count show an estimated NPV of 97% (PPV 69% with, 66% without *APOE4* allele count). Conversely, at high prevalences of PET-positivity (65%, 80%) the model predictors with and without *APOE4* allele count would have high PPVs estimates (94%-97%) but lower NPVs (68%-82%, **Table 3**).

**Table 3:** PPV and NPV for each model at different prevalences.

| Predictor | ptau-217 + Aβ42/40 |  | ptau-217 + Aβ42/40 +APOE4 allele count |  |
| --- | --- | --- | --- | --- |
| Prevalence | PPV (95% CI) | NPV (95% CI) | PPV (95% CI) | NPV (95% CI) |
| 20%, as expected in a primary care setting | 66% (54% to 79%) | 97% (95% to 98%) | 69% (56% to 80%) | 97% (95% to 99%) |
| 46%, observed study prevalence | 87% (78% to 93%) | 91% (83% to 95%) | 88% (80% to 93%) | 91% (84% to 95%) |
| 50%, as expected in MCI population | 89% (82% to 94%) | 89% (82% to 94%) | 90% (84% to 94%) | 90% (83% to 95%) |
| 65%, as expected in a general neurology setting | 94% (90% to 96%) | 82% (72% to 90%) | 94% (91% to 97%) | 82% (73% to 90%) |
| 80%, as expected in a memory care setting | 97% (95% to 98%) | 68% (54% to 81%) | 97% (95% to 98%) | 69% (55% to 81%) |

### 3.3 Stratification by cognitive status

Superimposing probability of PET positivity scores from the *APOE4* allele count model onto cognitive status showed significantly higher probability scores in AD individuals compared to MCI and NC (*P*<.001 for both), and significantly higher probability scores in MCI vs NC (*P*=0.002, **eFigure 1A**).

ROC-AUCs for independent model predictors for cognitive status were 0.943 (95%CI 0.894 to 0.992) for NC vs AD and 0.864 (95%CI 0.817 to 0.911) for MCI vs AD (**eFigure 1B**). However, the model predictor did not perform well by ROC analysis in distinguishing NC from MCI (AUC = 0.650, 95%CI 0.568 to 0.740).

### 3.4 Real-world data performance by age

For the 4,326 plasma specimens submitted to Quest Diagnostics for plasma Aβ42/40, ptau-217, and ApoE proteoform testing, the mean age of individuals was 73 years (SD 10 years, age range 28-98), most (58%) were female, and 38% had 1 or 2 *APOE4* alleles (**eTable 2**). In total, 42% (1,822) were classified as having a high likelihood, 51% (2,204) as having low likelihood, and 7% (300) as having an indeterminate likelihood for PET positivity (**Figure 3B**).

Based on their low likelihood for PET positivity, a PET scan or CSF testing would potentially be unnecessary for the 2,204 patients (**Table 4**). Similarly, imaging or CSF testing may not be required for the 1,822 high-likelihood patients based on the ≥90% sensitivity, specificity, PPV, NPV (**Table 3**) and recent guidance for a projected a prevalence of 50% of PET-positivity.^18^

**Table 4:** Proposed linear predictor cutpoints for clinical decision making.

| Probability of PET positivity range | Interpretation | Intended use population *N* (% of cohort) | PET positivity *N* (%) | MCI, *N* (%) | AD, *N* (%) | *APOE4*, *N* (%) | Real world, *N* (% of cohort) | Real world, projections of PET positivity, *N* (%)<sup>a</sup> | Follow-up evaluation and management considerations |
| --- | --- | --- | --- | --- | --- | --- | --- | --- | --- |
| < 0.3714 | Low likelihood | 101 (47.0%) | 9 (9.0%) | 101 (100%) | 0 (0%) | 16 (15.8%) | 2,204 (50.9%) | 198 (9.0%) | Consider non-AD etiology. |
| 0.3714 to 0.5375 | Indeterminant likelihood | 21 (9.8%) | 8 (38.1%) | 18 (85.7%) | 3 (14.3%) | 10 (47.6%) | 300 (6.9%) | 114 (38.0%) | Consider results within context of full differential; refer to a neurologist. |
| > 0.5375 | High likelihood | 93 (43.3%) | 82 (88.2%) | 51 (54.8%) | 42 (45.2%) | 56 (60.2%) | 1,822 (42.1%) | 1,607 (88.2%) | Consider results within context of full differential; refer to a neurologist; AD etiology likely. |
| All | All | 215 (100%) | 99 (46.0%) | 170 (79.1%) | 45 (20.9%) | 82 (38.1%) | 4,326 (100%) | 1,919 (44.4%) |  |

Abbreviations: AD, Alzheimer's disease; ApoE4, apolipoprotein E4 proteoform; MCI, mild cognitive impairment; PET, positron emission tomography.
<sup>a</sup>Real world projections of PET positivity are based on percent PET positivity in the intended use population cohort.

When parsed by the 3-biomarker model predictor designation that uses *APOE4* allele count, a significant difference in age between the low and high likelihood group (70.3 vs 75.5, ANOVA *P*<.001) and the low and indeterminate likelihood group (70.3 vs 76.3, ANOVA *P*<.001) was observed (**eTable 2**). No differences in age were seen between the indeterminate and high likelihood group.

Further stratifying the results by age, we observed a significant positive relationship between increasing median age and increasing probability of PET positivity (**eFigure 2**). For low likelihood patients, the percentage who are likely PET negative decreases with age, with 90.2% of patients younger than 50 years predicted to be amyloid PET negative compared to 37.2% for those 90 years or older (**eTable 4**).

### 3.5 Assay technical robustness and effects of bias and imprecision on likelihood reclassification

We evaluated the effects of total error, composed of analytical bias (technical error) and imprecision (random error), on likelihood reclassification when using the model predictor with *APOE4* allele count. In the current study, measured bias was −1.1% (95%CI, −3.1% to 0.9%) for ptau-217. Previously measured bias was 12% for Aβ42 and 11% for Aβ40.^8^ In the current study, the average measured imprecision for ptau-217, determined by taking the average of 2 QCs analyzed in duplicate over 14 nonconsecutive days, was 6%. The average imprecision for Aβ42/40, previously measured by taking the average imprecision across 5 study samples analyzed in duplicate over 25 days, was also 6%.^8,9^

Reclassification rates are shown in **eTable 5** and visual representation of classification comparisons with and without total error applied are shown in **eFigure 3**. Median reclassification after total error was applied across 10,000 simulations was 13.5% (9.8% to 16.7%). Within these data, most reclassifications were from indeterminate-to-negative: 3.3% (1.4% to 5.6%) and fewest were from negative-to-positive: 0.9% (0% to 2.3%).

## 4. DISCUSSION

The need for affordable, scalable, and accessible blood-based testing for Alzheimer’s disease has never been more important than with the emergence of the first disease-modifying therapies. We demonstrate that the combination of plasma Aβ42/40, ptau-217, and ApoE proteoform test results, measured using high-throughput LC-MS/MS and immunoassay methods, can predict the likelihood of amyloid pathology using an optimized model and a 2-cutpoint approach (**Table 2**). Automated specimen preparation and analysis allows a 96-well plate to be run in 2 to 8 hours, depending on the assay.

### 4.1 Model performance

The cutpoints were optimized for detecting amyloid PET status in patients with MCI and AD, who represent the intended-use population. In addition, these optimized cutpoints are in alignment with recently published performance recommendations to minimize the number of indeterminate results to ≤20% while maintaining a sensitivity and specificity of ≥90%.^18^ This ensures that our test can be used to both rule-in and rule-out amyloid PET positivity consistent with AD pathology as the likely culprit of cognitive impairment. The model minimized the proportion of indeterminate results to ≤10%, thus limiting results that can be uninformative to both patients and clinicians.

### 4.2 Model development

The optimized model was derived from logistic regression combining Aβ42/40, natural log transformed ptau-217 concentrations, and *APOE4* allele count. Each component provided a statistically significant contribution to the risk score, except for the homozygous *APOE4* component. All 12 homozygous *APOE4* individuals in our cohort were PET positive, and the absence of *APOE4* homozygotes in the PET-negative group provided no statistical control.

*APOE4* homozygosity has a prevalence of approximately 2% in the general population.^31^ We found homozygosity to be slightly higher both in our intended-use cohort and in clinical specimens (about 6%). However, *APOE4* homozygosity combined with PET-negativity would be expected to be extremely rare. For example, Fortea et al found almost 99% of *APOE4* homozygotes had some Alzheimer’s disease neuropathologic change.^32^ The same study suggested that *APOE4* homozygosity is now being considered a genetically distinct form of Alzheimer’s disease.^32^

The optimized model was slightly improved over Aβ42/40 combined with ptau-217 in terms of ROC-AUC and had lowest AIC score of all models (**Table 2**). Most importantly, the model substantially reduced the number of participants with an indeterminate result from 15% to 10%, with about two-thirds of the reclassified participants having a result more predictive of their actual PET status.

### 4.3 Comparisons with other models

The performance of combinations of biomarkers in predicting amyloid PET status has been reported by several different groups,^14–17^ including a study by Meyer et al^14^ who reported high predictive performance for a high-resolution MS-based assay incorporating ptau-217 normalized to non-phosphorylated tau (% ptau-217, ptau-217/np-tau-217) and Aβ42/40, optimized by a single cutpoint. The combined cohort used to develop the model in this study was less ethnically diverse than ours, although sub cohort analysis indicated that their model had equivalent accuracy regardless of age, sex, race, ethnicity, or *APOE4* status.

In contrast to the study by Meyer et al, the design of our study incorporated PET-positive and PET-negative participants, who were age-, sex-, ethnicity-, and years-of-education matched. While they reported on model robustness for subgroups, Meyer et al did not report on assay technical robustness, including the effects of technical and random error on patient classification. Our robustness analysis, which measured total error for both the Aβ42/40 and ptau-217 assays, demonstrated an overall reclassification rate of <14%, and just over 3% for the most risk reclassifications between 2 categories.

Other studies showing high performance using fewer biomarkers established PET-positivity using quantitative centiloid (CL) values with cutoffs of >25 CL or >24 CL indicating PET positivity. In contrast, we established PET positivity by central visual read,^10,14,16^ which is widely recognized as being highly accurate and the standard to which CL quantification is compared. Studies have demonstrated a 92% agreement between visual read and CL values. However, CL values were considered supportive in only 70% of cases that were considered challenging (11% of scans, expected rate in a memory clinic setting).^33^ Results suggest that CL values can be supportive when added to visual read assessment but have not necessarily been shown to be superior to the highly accurate visual read approach, with the authors acknowledging that “quantification should always be done in conjunction with visual assessment to avoid misclassifications due to potential quantification errors.” In addition, the authors note the recent adoption of opinion from the Committee for Medicinal Products for Human Use (EMACDOC-17005198181200791) stating that a “threshold of greater than 30 is reflective of established amyloid pathology at the individual level with high certainty.”^33^

### 4.3 Real-world data

Our real-world data set was composed of 4,326 consecutively run Aβ42/40, ptau-217, and ApoE proteoform specimens, submitted by healthcare providers, presumably for suspected dementia due to Alzheimer’s disease. As expected, we observed an increase in the likelihood of PET positivity with age, with predicted PET positivity rates ranging from 9% for individuals under the age of 50 to 55% for those 90 or older (**eTable 4**). These results suggest that PPV/NPV could be influenced by age-dependent trends in the prevalence of amyloid PET positivity, particularly for individuals under the age of 70.

Using our fixed cutpoints to categorize individuals into low, indeterminate, and high likelihood for amyloid PET positivity, we suggest that results may help guide physicians in clinical decision making (**Table 4**). Projecting the proportion of PET positivity from the intended-use population onto the 4,326 clinical specimens could potentially eliminate the need for PET or CSF testing on 2,204 individuals who are likely to have non-AD etiology, or slightly over 50% of tested individuals. Considering the cost of an amyloid PET scan in the United States (∼$5,000 USD),^34^ removing these individuals alone may potentially save $11,020,000 (less the cost of testing) as well as eliminating travel and wait times to receive imaging services.

### 4.3 Strengths and limitations

Key strengths of this study include its use of scalable, cost-effective plasma biomarkers in a cohort of ethnically diverse patients that are representative of the intended-use population. However, the models were developed using a cohort of participants in a memory care setting, which may not reflect the general clinical population. This concern is alleviated by similarity in the distribution between our intended-use cohort and the “real world” clinical specimens across the 3 risk classifications (**Figure 3**).

In addition, our study was not designed to evaluate the relevance or performance of these markers in the context of other causes of dementia, including mixed dementia with amyloid pathology or early onset Alzheimer’s disease or to address the effects of ethnically relevant genetic markers like *APOE4* on predictive performance of the tests. Also, the model will falsely identify PET-negative *APOE4* homozygote individuals as PET-positives, although these individuals are expected to be extremely rare. In our expanded cohort, we found one PET-negative *APOE4* homozygote individual, but they had normal cognition, excluding them from the intended-use population on which the model was built and predictors were trained.

Notably, *APOE4* status is also leading contributor to risk of amyloid-related imaging abnormalities (ARIA) with anti-amyloid therapies, however *APOE4* was included solely as a predictor of amyloid status in this study and results do not provide a quantitative measure of ARIA risk.^35^

Another limitation is the potential for high false positive rates that may be observed in a primary care setting (eg, PPV 69%) with low amyloid prevalence (eg, 20%). However, in such settings, the clinical utility of the models is better suited to rule-out AD pathology, given the high NPV’s (eg, 97%) observed in these low-prevalence populations.

Finally, while the model works well for predicting PET status and differentiates individuals with different cognitive status, models for predicting cognitive status only work well for differentiating NC from AD.

Replication of these results in other ethnically and socioeconomically diverse groups will support using these biomarkers for clinical assessment of potential candidates for anti-amyloid therapies. The combination of plasma biomarkers described herein is likely to complement clinical and radiologic diagnostic evaluations to further enhance the accuracy of AD diagnosis, especially when incorporated into enhanced models. For example, combining these models with assessments of cognitive function may offer a low-cost enhancement of model performance. Combining these models with specific imaging methodology to assess diverse neurodegenerative processes may be important in selecting patients who are most likely to respond to specific therapeutic interventions.^36^

## Data Availability

The data supporting the conclusions of this article will be made available by the authors, without undue reservation.

## Study Funding

Funding for this work was provided by NIH Research Grants R01NS052318 and R01NS075012 (D.E. Vaillancourt).

## Disclosures

D.M. Weber is an employee of Quest Diagnostics. He also holds patents for the detection of beta-amyloid by mass spectrometry as well as for the detection of apolipoprotein E proteoforms by mass spectrometry; M.A. Stroh is an employee of Quest Diagnostics; S.W. Taylor is an employee of Quest Diagnostics; R.J. Lagier is an employee of Quest Diagnostics; J.Z. Louie is an employee of Quest Diagnostics; N.J. Clarke is an employee of Quest Diagnostics. He also holds patents for the detection of beta-amyloid by mass spectrometry as well as for the detection of apolipoprotein E proteoforms by mass spectrometry; D.E. Vaillancourt reports no disclosures relevant to the manuscript; S. Rayaprolu reports no disclosures relevant to the manuscript; R. Duara reports no disclosures relevant to the manuscript; M.K. Racke is an employee of Quest Diagnostics.

## Notes

### Competing Interest Statement

DW, MS, ST, RL, JL, NC, and MR are employees of Quest Diagnostics. DW and NC hold patents for the detection of beta-amyloid by mass spectrometry as well as for the detection of apolipoprotein E proteoforms by mass spectrometry. The remaining authors declare that the research was conducted in the absence of any commercial or financial relationships that could be construed as a potential conflict of interest.

### Funding Statement

Funding for this work was provided by National Institutes of Health (NIH) Center Core Grant P30AG066506 (TEG) and NIH Research Grants R01NS052318 and R01NS075012 (DEV).

### Author Declarations

The study was approved by the Mount Sinai Medical Center IRB, and all participants provided informed consent.

### Summary of Updates

This manuscript has been updated to include both ethnicity and race, as well as a breakdown of Hispanic origin, for the study cohort.

