## Supplemental Material for "Development and clinical validation of blood-based multibiomarker models for the evaluation of brain amyloid pathology"

**SUPPLEMENTARY MATERIAL**

**Supplementary Figures**

**eFigure 1**: A) Cognitive status superimposed on cutpoints developed using 3-marker model predictor for PET-positivity that includes *APOE4* allele counts. Amyloid PET-negative individuals are illustrated with orange circles and amyloid PET-positive individuals are illustrated with blue circles. Horizontal dashed lines show cutpoints fixed to achieve 91% sensitivity and 91% specificity. B) Independent ROC-AUC analysis for prediction of cognitive outcomes (also including *APOE4* allele count). Abbreviations: AD, Alzheimer’s disease; NC, normal cognition; MCI, mild cognitive impairment

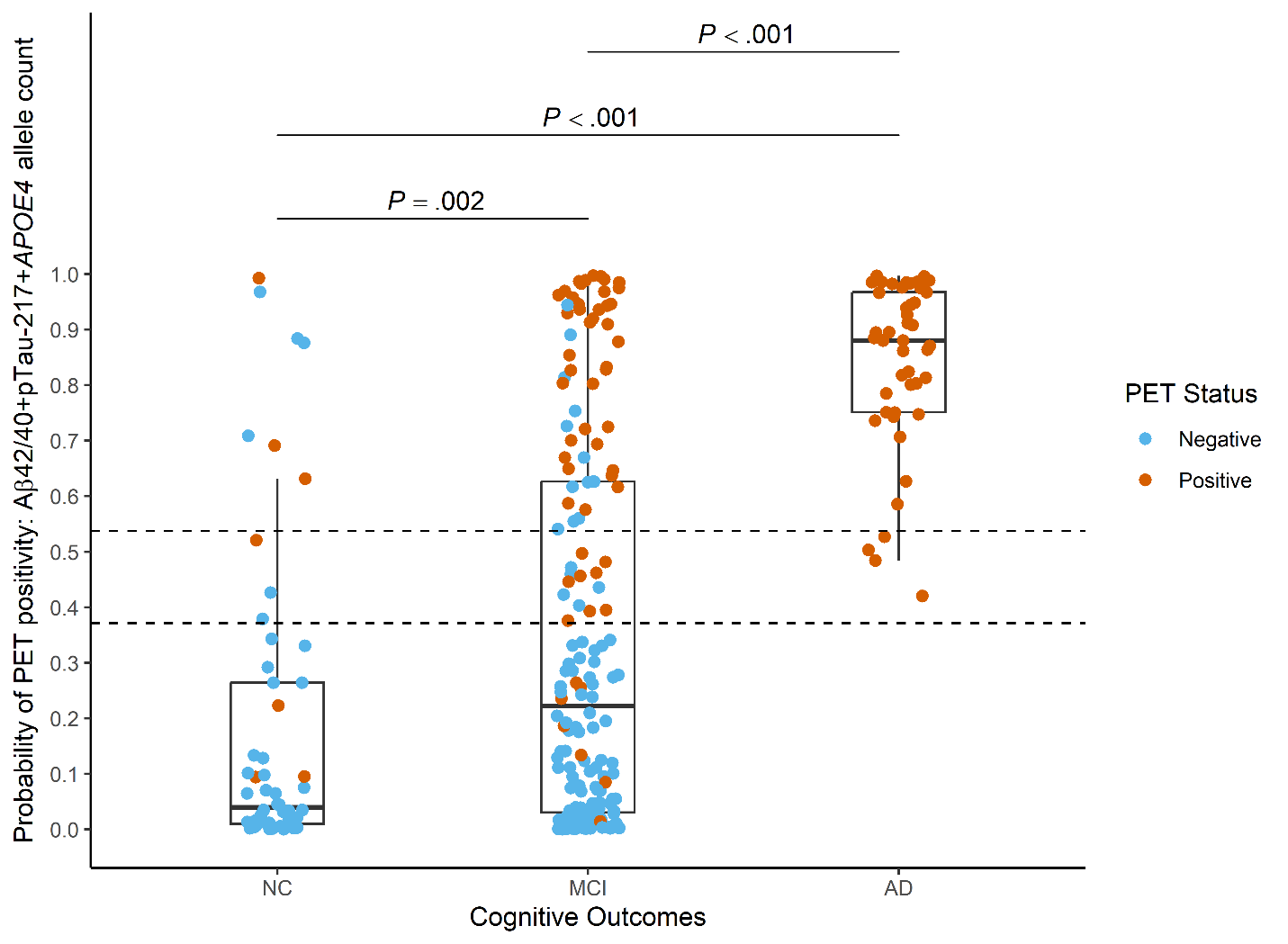

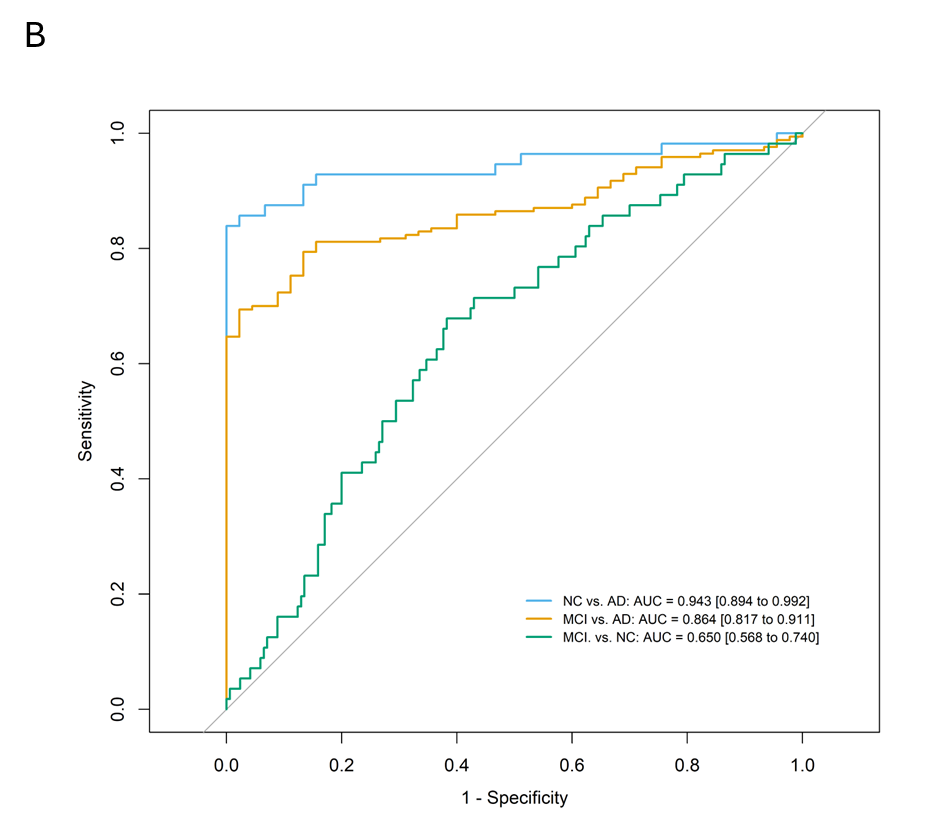

**eFigure 2**: Distribution of model predictor probability scores by age for the 4,326 real-world specimens. Horizontal dashed lines show cutpoints fixed to achieve 91% sensitivity and 91% specificity. A significant positive relationship between increasing median age and increasing probability of PET positivity (Spearman’s rho 0.279, 95% CI = 0.251 to 0.307, *P* < .001) was observed.

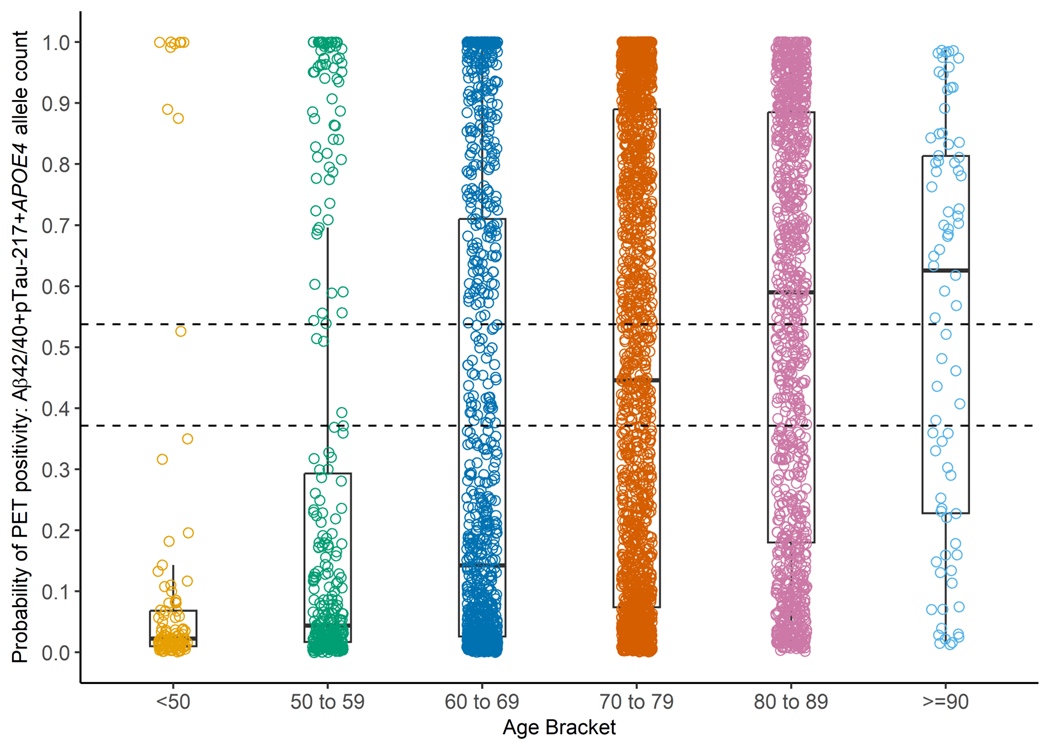

**eFigure 3:** Median reclassification of with total error applied over 10,000 simulations. Dashed lines are probability score cutpoints determined by the 3-marker model that includes *APOE4* allele count.

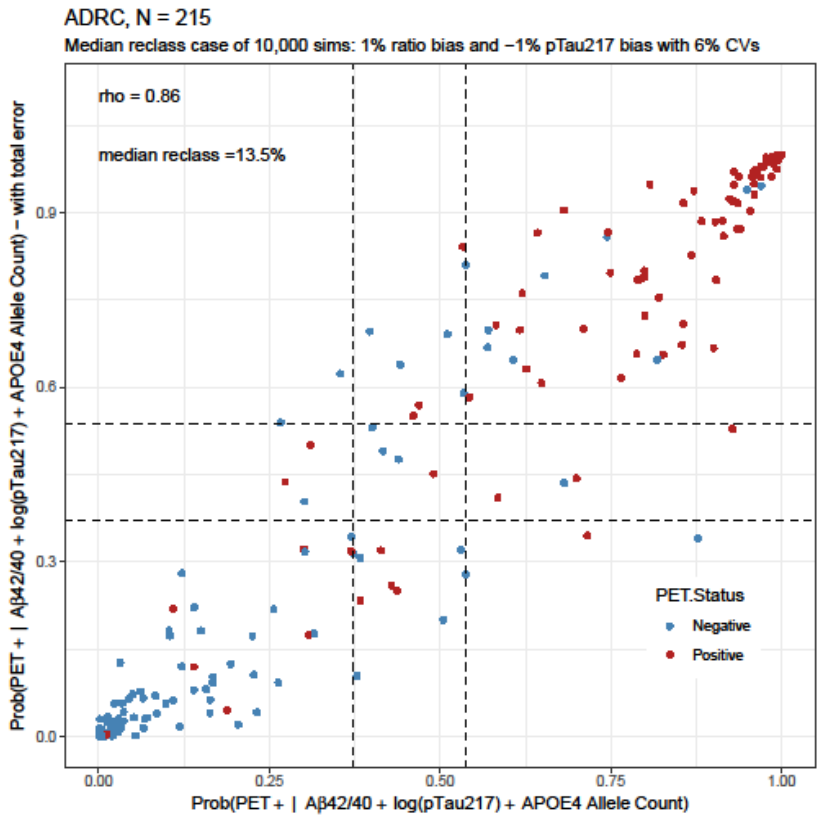

**Supplementary Tables**

**eTable 1:** Participant characteristics by cognitive status

| **Characteristic** | **Full Cohort [N=271]** | **Normal Cognition (NC) [N=56]** | **Mild Cognitive Impairment (MCI) [N=170]** | **AD [N=45]** | ***p*-value^a^** |
| --- | --- | --- | --- | --- | --- |
| **Demographics** | | | | |  |
| **Age** | | | | |  |
| Mean (SD) | 78.4 (8.4) | 76.4 (8.1) | 79.3 (8.2) | 77.5 (9.2) | 0.060 |
| **Sex** | | | | |  |
| % female [N] | 60.1% [163] | 75.0% [42] | 54.7% [93] | 62.2% [28] | *0.024* |
| **Ethnicity** | | | | |  |
| % Hispanic [N] | 52.8% [143] | 46.4% [26] | 54.7% [93] | 53.3% [24] | 0.574 |
| Central American | 7 | 0 | 6 | 1 |  |
| Cuban | 78 | 13 | 52 | 13 |  |
| Dominican | 1 | 0 | 0 | 1 |  |
| Puerto Rican | 10 | 1 | 5 | 4 |  |
| South American | 46 | 12 | 29 | 5 |  |
| Missing | 1 | 0 | 1 | 0 |  |
| **Race: Hispanic** | | | | |  |
| Asian | 0 | 0 | 0 | 0 |  |
| Black or African American | 2 | 1 | 1 | 0 |  |
| Multiracial | 1 | 0 | 1 | 0 |  |
| White | 139 | 25 | 90 | 24 |  |
| Missing | 1 | 0 | 1 | 0 |  |
| **Race: Non-Hispanic** | | | | |  |
| Asian | 1 | 1 | 0 | 0 |  |
| Black or African American | 10 | 1 | 7 | 2 |  |
| Multiracial | 1 | 1 | 0 | 0 |  |
| White | 111 | 25 | 69 | 17 |  |
| Missing | 5 | 2 | 1 | 2 |  |
| **Education** | | | | |  |
| Mean years (SD) | 15.3 (3.2) | 16.0 (2.9) | 15.3 (3.2) | 14.5 (3.5) | 0.080 |
| **Cognitive Assessment** | | | | |  |
| **MMSE** | | | | |  |
| Median (Interquartile) | 28 (25 to 30) | 29 (29 to 30) | 28 (27 to 29) | 21 (17 to 24) | *< 0.001* |
| **CDR** | | | | |  |
| % CDR > 0 [N] | 77.8% [211] | 14.2% [8] | 92.9% [158] | 100% [45] | *< 0.001* |
| ***APOE4*** | | | | |  |
| % *APOE4+* carriers [N] | 35.0% [95] | 23.2% [13] | 33.5% [57] | 55.6% [25] | *0.003* |
| %*APOE4* allele count 1 [N] | 30.0% [81] | 19.6% [11] | 28.2% [48] | 88.0% [22] |  |
| %*APOE4* allele count 2 [N] | 5.2% [14] | 3.6% [2] | 5.3% [9] | 6.7% [3] |  |
| **Plasma Biomarkers** | | | | |  |
| **Aβ42, pg/mL** | | | | |  |
| Mean (SD) | 61.2 (17.6) | 63.9 (14.9) | 62.9 (18.3) | 51.1 (14.6) | *< 0.001* |
| **Aβ40, pg/mL** | | | | |  |
| Mean (SD) | 379.5 (84.0) | 365.2 (67.1) | 385.6 (86.2) | 373.9 (93.2) | 0.260 |
| **Aβ42/40** | | | | |  |
| Mean (SD) | 0.161 (0.029) | 0.176 (0.028) | 0.163 (0.030) | 0.136 (0.015) | *< 0.001* |
| **ptau-217, pg/mL** | | | | |  |
| Median (Interquartile) | 0.21 (0.09 to 0.51) | 0.11 (0.06 to 0.26) | 0.17 (0.09 to 0.40) | 0.67 (0.44 to 0.97) | *< 0.001* |
| **Model with *APOE4* allele count** | | | | |  |
| Median (Interquartile) | 0.3004 (0.0304 to 0.7534) | 0.0400 (0.0102 to 0.2643) | 0.2227 (0.0304 to 0.6263) | 0.8803 (0.7513 to 0.9675) | *< 0.001* |

Abbreviations: Aβ, beta-amyloid; AD, Alzheimer’s disease *APOE4*, APOE ε4; CDR-SB, clinical dementia rating sum of boxes; MMSE, mini-mental state examination

^a^ *P*-values are derived from ANOVA or Kruskal-Wallis test for continuous variables, and Fisher's exact test for categorical variables between NC, MCI, and AD subgroups. *P*-values < 0.05 are highlighted in italics.

**eTable 2**: Demographics for the 4,326 real-world specimens submitted for plasma Aβ42/40, ptau-217, and ApoE proteoform testing

| **Characteristic** | **Total cohort [N = 4326]** | **Low likelihood [N = 2204]** | **Indeterminate likelihood [N = 300]** | **High likelihood [N = 1822]** |
| --- | --- | --- | --- | --- |
| **Age** | | | | |
| Mean (SD) | 72.9 (9.8) | 70.3 (10.6)^a,b^ | 76.3 (7.5)^b^ | 75.5 (8.2)^a^ |
| **Sex** | | | | |
| % female [N] | 57.7% [2497] | 56.9% [1253] | 52.7% [158] | 59.6% [1086] |
| **Model with *APOE4* allele count** | | | | |
| Mean (SD) | 0.4396 (0.3803) | 0.0969 (0.0992) | 0.4551 (0.0482) | 0.8513 (0.1390) |
| **Plasma Aβ42/40** | | | | |
| Mean (SD) | 0.148 (0.019) | 0.158 (0.017) | 0.144 (0.014) | 0.137 (0.015) |
| **Plasma Aβ42 (pg/mL)** | | | | |
| Mean (SD) | 49.1 (13.5) | 49.5 (11.1) | 49.8 (13.9) | 48.5 (15.2) |
| **Plasma Aβ40 (pg/mL)** | | | | |
| Mean (SD) | 333.7 (97.7) | 313.7 (69.6) | 345.3 (86.8) | 356.1 (120.6) |
| **Plasma ptau-217 (pg/mL)** | | | | |
| Median (IQR) | 0.17 (0.08 to 0.39) | 0.08 (0.06 to 0.13) | 0.23 (0.15 to 0.29) | 0.44 (0.29 to 0.71) |
| ***APOE4*** | | | | |
| %*APOE4*+ carriers [N] | 37.8% [1635] | 15.5% [341] | 33.0% [99] | 65.6% [1195] |
| %*APOE4* allele count 1 [N] | 32% [1386] | 15.5% [341] | 33.0% [99] | 52.0% [946] |
| %*APOE4* allele count 2 [N] | 5.8% [249] | 0% [0] | 0% [0] | 13.7% [249] |

^a^ ANOVA *P* < .001

^b^ ANOVA *P* < .001

Abbreviations: Aβ, beta-amyloid; *APOE4*, *APOE* ε4.

**eTable 3:** Model predictor performance at a single cutpoint for individuals and combined biomarkers. Youden index was used to identify cutpoints that maximize sensitivity and specificity for each evaluated model.

| **Predictor: Single cutpoint** | **Optimal cutpoint** | | **AUC (95% CI)** | **Sensitivity (95% CI)** | **Specificity (95% CI)** | **Accuracy** | **AIC** |
| --- | --- | --- | --- | --- | --- | --- | --- |
| ptau-217, pg/mL | | 0.2300 | 0.846  (0.808 to 0.905) | 87%  (77% to 92%) | 72% (64% to 80%) | 79% (73% to 84%) | 263 |
| Aβ42/40, ratio | | 0.1600 | 0.851  (0.796 to 0.905) | 93%  (86% to 97%) | 74% (66% to 81%) | 83% (77% to 87%) | 256 |
| Model 1: ptau-217+Aβ42/40, probability score | | 0.4233 | 0.929  (0.894 to 0.963) | 87% (79% to 92%) | 84% (76% to 89%) | 85% (80% to 89%) | 195 |
| Model 2: ptau217 + Aβ42/40 + *APOE4* carrier status, probability score | | 0.6181 | 0.938  (0.906 to 0.969) | 81% (72% to 87%) | 94% (88% to 97%) | 88% (83% to 92%) | 189 |
| Model 3: ptau217 + Aβ42/40 + *APOE4* allele count, probability score | | 0.5761 | 0.942  (0.912 to 0.972) | 82% (73% to 88%) | 93% (87% to 97%) | 88% (83% to 92%) | 186 |

Abbreviations: Aβ, beta-amyloid; AIC, Akaike Information Criterion; APOE4, apolipoprotein E ε4; AUC, area under the curve; CI, confidence interval.

**eTable 4**: Demographics for the 4,326 real-world specimens submitted for plasma Aβ42/40, ptau-217, and ApoE proteoform testing parsed by decade of age.

| **Age Group, years** | **% [N]** | **Probability of PET positivity score median [interquartile]** | **Probability of PET positivity designation** | | |
| --- | --- | --- | --- | --- | --- |
|  |  |  | **Low likelihood, % [N]** | **Indeterminate likelihood, % [N]** | **High likelihood, % [N]** |
| **< 50^a^** | 2.4% [102] | 0.0227 [0.0101 to 0.0679] | 90.2% [92] | 0.9% [1] | 8.8% [9] |
| **50 to 59** | 7.5% [323] | 0.0437 [0.0168 to 0.2928] | 77.1% [249] | 1.2% [4] | 21.7% [70] |
| **60 to 69** | 21.4% [926] | 0.1427 [0.0256 to 0.7100] | 63.8% [591] | 4.8% [44] | 31.4% [291] |
| **70 to 79** | 42.8% [1,852] | 0.4456 [0.0737 to 0.8901] | 46.3% [858] | 7.6% [141] | 46.1% [853] |
| **80 to 89** | 24.2% [1,045] | 0.5900 [0.1804 to 0.8853] | 36.8% [385] | 10.0% [104] | 53.2% [556] |
| **≥ 90^b^** | 1.8% [78] | 0.6252 [0.2281 to 0.8135] | 37.2% [29] | 7.7% [6] | 55.1% [43] |
| **All** | 100% [4,326] | 0.3495 [0.0526 to 0.8462] | 50.9% [2,204] | 6.9% [300] | 42.1% [1,822] |

^a^ Age range: 28 to 50 years old

^b^ Age range: 90 to 98 years old

Abbreviations: PET, positron emission tomography

**eTable 5:** Simulated reclassification rates when using the 3-marker model including *APOE*4 allele count for total error (TE) (N = 215)

| **Comparison** | **Median reclassification** |
| --- | --- |
| **Overall reclassification rate** | **13.5% (9.8% to 16.7%)** |
| Negative to Indeterminate | 1.8% (0.5% to 3.7%) |
| Negative to Positive | 0.9% (0% to 2.3%) |
| Indeterminate to Negative | 3.3% (1.4% to 5.6%) |
| Indeterminate to Positive | 2.8% (0.9% to 4.6%) |
| Positive to Indeterminate | 2.8% (0.9% to 4.6%) |
| Positive to Negative | 1.4% (0% to 2.8%) |
